# Feasibility of FFR-CT and CCTA features to predict the hemodynamic changes of high take-off coronary artery

**DOI:** 10.1101/2023.08.24.23294591

**Authors:** Jiong Ni, Shuihua Cheng, Peijun Wang, Chenjin Ge

## Abstract

**Background:** High take-off coronary artery (HTCA) is a neglected coronary artery anomaly with debated clinical importance. Determining the pathophysiological significance of this anomaly remains challenging. The aim of this study was to explore clinical symptoms, anatomic features based on CCTA and FFR-CT which will help to predict significant hemodynamic change of HTCA.

**Methods:** This retrospective study recruited 72 patients with HTCA undergoing CCTA and invasive coronary angiography. Demographic, clinical characteristics, anatomic and functional features based on CCTA were collected to identified independent risk factors by multivariate logistical analysis. Receiver operating characteristic curve analysis was performed to determine the predictive accuracy of these factors.

**Results:** Chest pain and the angle at the HTCA were identified as independent risk factors (p=0.005, p=0.002) associated hemodynamical significant HTCA. The value of FFR-CT was consistency with FFR in HTCA patients (VIF=73.811). The index of FFR-CT had significantly larger AUC than the index of chest pain (p=0.003) and similar AUC with the angel at the HTCA (p=0.059). Compared with combined index of angle at HTCA plus chest pain, FFR-CT had a larger AUC (p=0.000). The similar AUC was found between FFR-CT and the combined index of chest pain, angle at the HTCA plus FFR-CT (p=0.2359).

**Conclusions:** FFR-CT, chest pain and angel at HTCA based on CCTA played a great role to predict significant hemodynamic changes of HTCA. FFR-CT had a superior diagnostic performance to predict abnormal hemodynamic changes in patients with HTCA.

## Introduction

High take-off coronary artery (HTCA) is abbreviated as take-off coronary artery, referring to the origin of either the right coronary artery (RCA) or the left coronary artery (LCA) arising from the ascending aorta [1]. HTCA was reported in 0.20% of the general population, with RCA accounting for approximately 84% of all HTCA reported [2]. It is usually a neglected coronary artery anomaly. HTCA may occur alone or in association with other anomalies, and its clinical importance has been debated. Most of this anomaly is benign without clinical problems. However, in real world, the cases of ischemia, myocardial infarction, syncope and sudden cardiac death were continuously reported in HTCA [3-5], raising clinical concerns. HTCA is also considered as a risk factor for ischemia without atherosclerosis [2]. Actually, determining the pathophysiological significance of this anomaly remains a challenge, especially in isolated HTCA. Wang et al. reported that the symptoms of a HTCA were associated with the site and its course of progression [6]. Loukas et al. believed that the height of the HTCA and the acute angle at the HTCA indicated clinical importance [2]. A distance between the origin and sinus equal to or greater than 10 mm was thought to be clinically important [2]. These inferred HTCA with significant hemodynamic changes may be caused by many risk factors, and combination of these risk factors might play a role in the development of HTCA.

Assessment of hemodynamic significance is important to guide therapy in patients with HTCA. Although fractional flow reserve (FFR) is a gold standard in evaluating hemodynamical significant in coronary stenoses, it was limited to be used in coronary anomaly. The main deficit was that FFR was an invasive method, it was not a proper method to be used in most benign hemodynamic changes [7]. With the development of multidetector computed tomography, coronary computed tomography angiography (CCTA) has shown superiority in delineating the origin and proximal path of anomalous coronary arteries [8]. CT-based fractional flow reserve (FFR-CT) simulation, with a value ≤ 0.8 indicating the presence of a coronary lesion, offers a functional evaluation of coronary disease, including coronary artery anomaly [9-10]. Moreover, FFR-CT provides better diagnostic accuracy than CCTA alone for predicting hemodynamically significant lesions [10-11].

Therefore, the goal of this study was to explore clinical symptoms, anatomic features based on CCTA and FFR-CT which will help to predict significant hemodynamic change of HTCA.

## Methods

### Patient selection

This retrospective anonymous study was approved by the Ethics Committee and the Ethics Committee waived the need for informed consent. Data from a total of 19,331 patients who had undergone CCTA at two hospitals between January 2017 and December 2021 were retrospectively analyzed. 119 patients (0.62%) with HTCA were found in CCTA. Patients with abnormal EKG, like T waves changes, ST segment elevation or depression, or A-V blocks were elected to undergo further invasive coronary angiography within 1-2 weeks after CCTA. Among these patients, patient with a history of valvopathies, congenital heart diseases and coronary artery bypass grafting were first excluded. Besides these, patients who had one of the following were excluded from this study: (1) a coronary plaque and at least one lesion with ≥50% stenosis on major epicardia vessels (diameter ≥2 mm); (2) poor quality of CCTA images (defined as images with moderate or severe artifact, Likert scale score 2 and score 1) [12]; (3) patients with diffuse calcified plaques. Registered patients were divided into two groups based on the value of FFR: the FFR ≤ 0.8 group, a critical cutoff value indicating abnormal hemodynamics, and the FFR > 0.8 group, indicating normal hemodynamics [10, 13].

### Data collection

Demographic and baseline clinical characteristics including age, gender, previous medical history and clinical manifestations of all patients were collected from medical records and CCTA reports. Since ischemia is a risk of this anomaly, previous related histories and clinical symptoms were collected. Hypertension, diabetes mellitus and dyslipidemia were recorded as previous history in this study. Chest pain (including angina-like chest pain, atypical angina-like chest pain and chest discomfort, that were scored as 1, 2, and 3), syncope, dyspnea, and palpitation were recorded as clinical symptoms [8].

### Imaging protocol of CCTA

CCTA was performed for all patients with a 320-slice multidetector CT (Aquillion One, Toshiba, Japan) or a 64-slice multidetector CT (Aquillion, Toshiba, Japan). All patients with a heartbeat > 70 beats per minute (bpm) were orally administered with a beta-blocker (25-50 mg, AstraZeneca pharmaceutical Limited Company, China) 30 minutes before CCTA examination. Prior to CCTA examination, the heart rate was reevaluated to ensure that it was < 70 bpm. A test bolus was first injected and the region of interest was placed on the ascending aorta to find the exact delay scan time, which was defined as the CT value at 200 Hounsfield units. Patients were then injected with a bolus contrast media (lopamidol, 370 iodine/ml, Bracco Sine, China) into the antecubital vein at a rate of 4.5-5 ml/s, followed by 20-40 ml saline flush. Patient’s weight and scanning time were used to determine the amount of contrast media required.

A 64-slice multidetector CT was used to perform retrospective electrocardiography (ECG)-gated CCTA with the reconstruction slice thickness=0.5 mm, the collimation= 64×0.625 mm, and the reconstruction slice interval=0.3 mm. The tube voltage was 120 Kv and the rotating time of the tube was 9.2s. The automatic tube current modulation (ATCM) technique was used with the noise index being set to 20. A 320-slice multidetector CT was used to perform prospective ECG-gated CCTA, with the set trigger window being set to 70%. The pitch and tube current were modified with the R-R interval and the effective current was 150 mAs. The full dose during the R-R interval and the effective current was 65%-80% below 70 bpm and 40%-85% above or equal to 70 bpm, respectively. The reconstruction slice interval was 0.5 mm, and the collimation was 320×0.5 mm. The parameters of the prospective acquisition and those of the retrospective acquisitions were the same. The dose length of the product was 404.73± 25.23 Gy.

### CCTA image reconstruction and analysis

Data was transferred to an offline workstation (Vitrea 4.1, Toshiba, Japan) for further analysis. Post-reconstruction and measurements were performed using images of cardiac phases with the least motion. Axial images, multiplanar reconstruction (MPR), curved planar reformation (CPR), maximum intensity projection (MIP), and volume rending (VR) images were available for evaluation. Two experienced cardiovascular radiologists with 16-years’ and 12-years’ experience in cardiovascular imaging respectively (who were blinded to clinical histories) independently analyzed these HTCA images.

HTCA was diagnosed based on CCTA examination if the origin of a coronary artery was 10 mm above coronary sinus at VR images. A HTCA has four subtypes: LCA above the left coronary sinus, LCA above the right coronary sinus, RCA above the right coronary sinus, and RCA above the left coronary sinus. The number of these four subtypes of HTCA was compared between FFR ≤ 0.8 and FFR > 0.8 groups. The height of the HTCA and the angle at the HTCA were considered as high-risk anomalous anatomical signs to be documented [14]. Furthermore, the distance between the origin of the HTCA and the vertical line of left/right commissure was taken as a reference point and measured separately in every area. The diameter midpoint in the origin of the HTCA was defined as a measuring point. The distance above the left coronary sinus was marked distance A and the distance above the right coronary sinus was marked distance B. These radiological findings were measured at both MPR and VR images. A HTCA with a plaque, intraarterial course (course between pulmonary trunk and aorta) and intramural course were observed at both the axial and CPR images.

### FFR-CT stimulation and measurement

The FFR-CT calculations were performed on the routine CCTA database using a software (version 3.0, United Imaging, China). The software was designed based on the 3D-CFD model using transluminal attenuation gradient algorithm [15]. Images of the phases of end diastole and end systole were used to create a coronary model. The boundary areas including inlet, outlet and blood wall were set. Grip processing was then used in the coronary model. Clinical data, such as blood pressure and heart rate, were obtained and used to adjust the boundary area. Material properties, fluid model, simulation parameters and convergence conditions were set at the same time. Once all of these were accepted, a color-coded coronary tree with corresponding FFR values was automatically generated [16].

### ICA and FFR measurement

Invasive coronary angiography (ICA) was performed and independently evaluated by two experienced interventional cardiologists (with 22-years’ and 18-years’ experience in coronary intervention, respectively), who were blinded to the results of FFR-CT. Visual assessment was used to determine the stenotic extent of each lesion based on at least two views for each major coronary artery. FFR measurement was performed using a 0.014-in. pressure guidewire (St Jude Medical) after hyperemia was successfully induced by intravenous infusion of adenosine at a dose of 140 μg/(kg min). Lesions with a FFR≤0.8 were considered a significant hemodynamical change.

### Statistical analysis

All statistical analysis was performed using SPSS version 23.0 (SPSS Inc, Chicago, USA) and R Statistical Software version 4.0.3 (R Foundation for Statistical Computing). One-sample Kolmogorov-Smirnov test was used to determine the normal distribution of data. Normally distributed variables were expressed as mean±standard deviation and abnormally distributed variables were expressed as median and quartiles. Student’s t-tests were used for comparison of normally distributed data while the Mann Whitney U-tests were used for abnormally distributed data. Categorical variables are expressed as a percentage (%), and compared with chi-square test and Fisher exact test. Bonferroni Z test was used to further analyze sub-categorical variables. Univariate logistic regression analysis was performed to identify predictive factors for all p values with two-sided evaluation and those factors with a p value < 0.10 identified in univariate analysis were included in a multivariate analysis. Multivariate logistic regression analysis was performed to identify independent predictive factors, and a stepwise method was used to identify the risk factors that could most accurately predict significant hemodynamic changes. Sensitivity, specificity, and area under the curve (AUC) of independent predictive factors were generated from receiver operated characters (ROC) curves. ROC curves with different factors and combined factors were compared. A p-value < 0.05 was considered statistically significant.

## Results

### Demographic and baseline clinical characteristics analysis between the two groups

A total of 119 cases presenting with a HTCA between January 2017 and December 2021 were initially included in this study. After the application of the exclusion criteria, a total of 72 patients (mean age: 47.54 ± 9.13 years; 40 females and 32 males) with isolated HTCA were finally enrolled in this study. These patients were divided into two groups according to the value of FFR: the FFR ≤ 0.8 group (n = 25) and the FFR > 0.8 group (n = 47) (Fig. 1).

**Figure 1.**
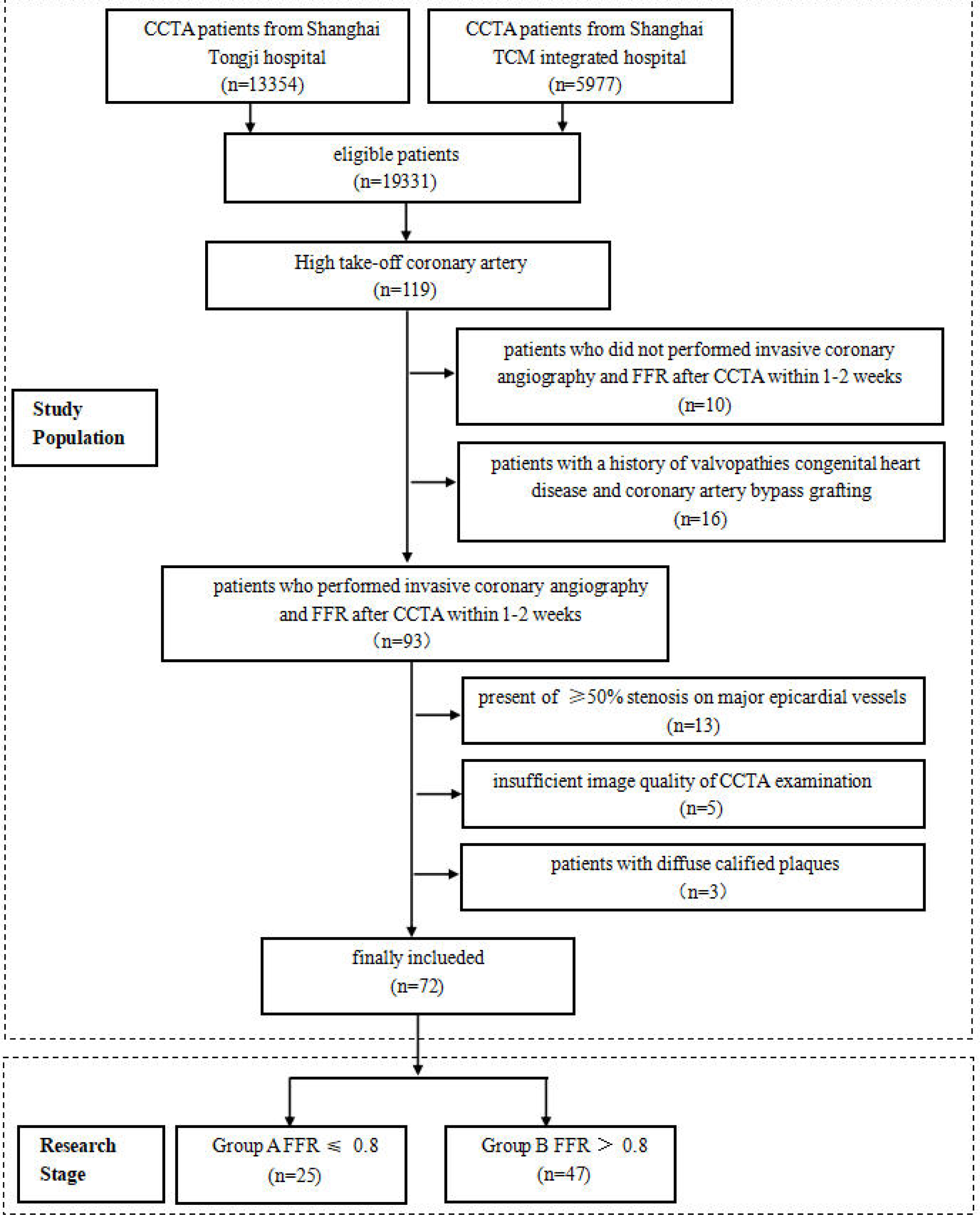
Flow chart shows the procedure of patient selection. Abbreviations: CCTA= Coronary computed tomography angiography, ICA=invasive coronary angiography, FFR= Fractional flow reserve

As shown in Table 1, there were no significant differences in gender (p= 0.956) or previous history between these two groups (p= 0.224, 0.502, 0.156). However, the FFR > 0.8 group were older than the FFR ≤ 0.8 group (p= 0.002).

**Table 1.**
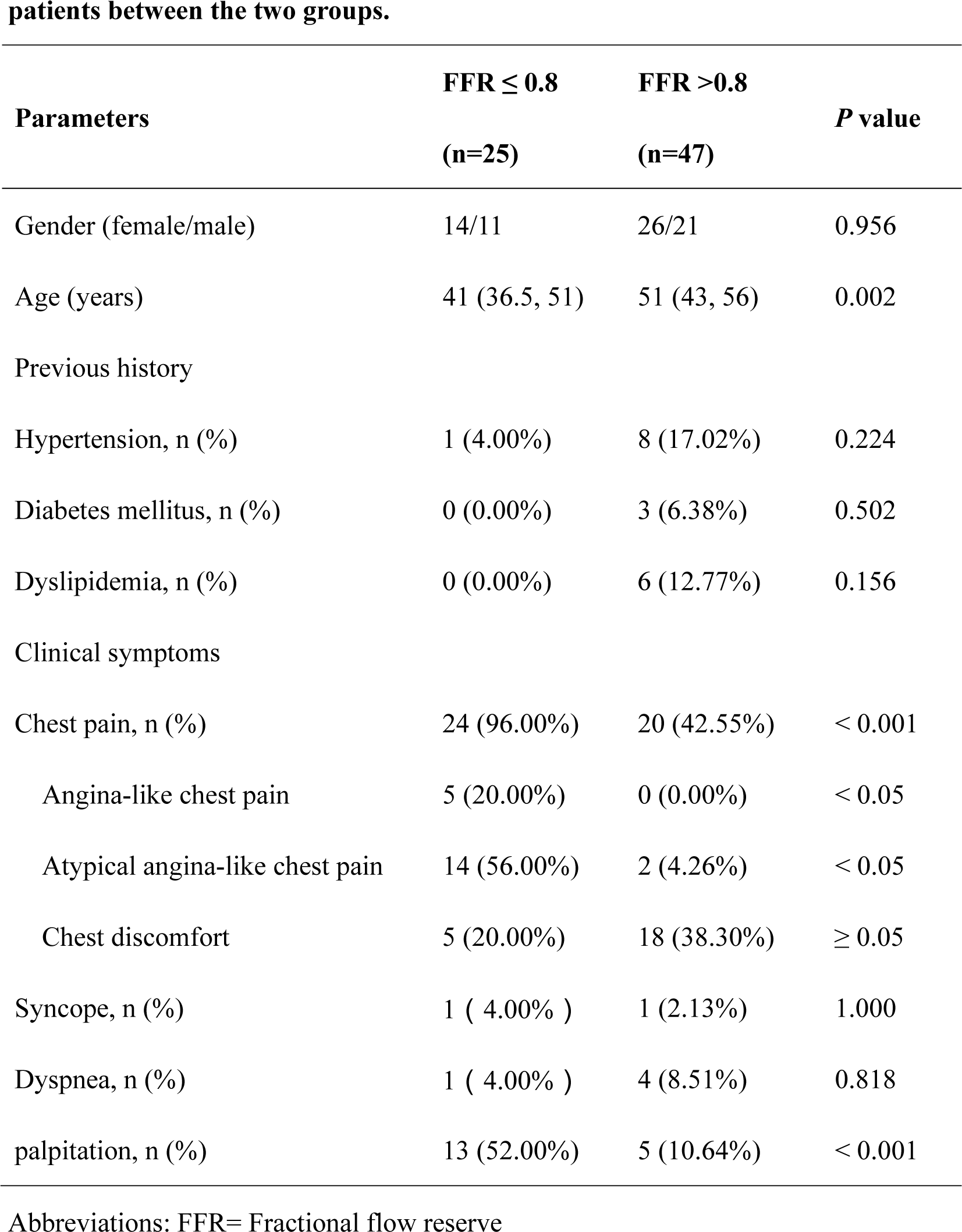
Comparison of demographic and baseline clinical characteristics of.

patients between the two groups.
| **Parameters** | **FFR ≤ 0.8**<br>**(n=25)** | **FFR $>$0.8**<br>**(n=47)** | **P value** |
| --- | --- | --- | --- |
| Gender (female/male) | 14/11 | 26/21 | 0.956 |
| Age (years) | 41 (36.5, 51) | 51 (43, 56) | 0.002 |
| Previous history |  |  |  |
| Hypertension, n (%) | 1 (4.00%) | 8 (17.02%) | 0.224 |
| Diabetes mellitus, n (%) | 0 (0.00%) | 3 (6.38%) | 0.502 |
| Dyslipidemia, n (%) | 0 (0.00%) | 6 (12.77%) | 0.156 |
| Clinical symptoms |  |  |  |
| Chest pain, n (%) | 24 (96.00%) | 20 (42.55%) | $< 0.001$ |
| Angina-like chest pain | 5 (20.00%) | 0 (0.00%) | $< 0.05$ |
| Atypical angina-like chest pain | 14 (56.00%) | 2 (4.26%) | $< 0.05$ |
| Chest discomfort | 5 (20.00%) | 18 (38.30%) | $\geq 0.05$ |
| Syncope, n (%) | 1 ( 4.00% ) | 1 (2.13%) | 1.000 |
| Dyspnea, n (%) | 1 ( 4.00% ) | 4 (8.51%) | 0.818 |
| palpitation, n (%) | 13 (52.00%) | 5 (10.64%) | $< 0.001$ |

Abbreviations: FFR= Fractional flow reserve

The FFR ≤ 0.8 group presented with more symptoms of chest pain (p <0.001) and palpitation (p<0.001) than in the FFR > 0.8 group. No significant difference was observed between groups with regard to the symptoms of syncope or dyspnea. The symptom of chest pain was subclassified into angina-like chest pain, atypical angina-like chest pain and chest discomfort. When these subclassifications were compared between the two groups, the FFR ≤ 0.8 group had more symptoms of angina-like chest pain, atypical angina-like chest pain.

### Anatomical features of HTCA and FFR-CT analysis between the two groups

Among these 72 patients, the number of LCA above the left coronary sinus was 13 (18.06%), the number of LCA above the right coronary sinus 14 (19.44%), the number of RCA above the right coronary sinus 30 (41.67%), and the number of RCA above the left coronary sinus 15 (20.83%). However, these subtypes of HTCA were not related with significant hemodynamic change (p= 0.742). Distance A (distance above the left coronary sinus) was measured in 43 patients and distance B (distance above the right coronary sinus) was measured in 29 patients. Distance A was 2.70 ± 1.49 mm and distance B was 4.55 ± 2.64 mm. The height of the HTCA was 13.71 ± 2.01 mm and the angle at the HTCA was 47.22 ± 17.63 degrees. Four patients had mixed plaque, nine had intraarterial course, and three patients had intermural course. As shown in Table 2, just angle at HTCA had a significant difference in the FFR ≤ 0.8 group compared with the FFR > 0.8 group (p <0.001). Representative cases are given in Fig. 2. The value of FFR-CT was consistency with FFR in HTCA patients (p<0.001). The Kenduall values were 0.918 (p<0.001).

**Table 2.**
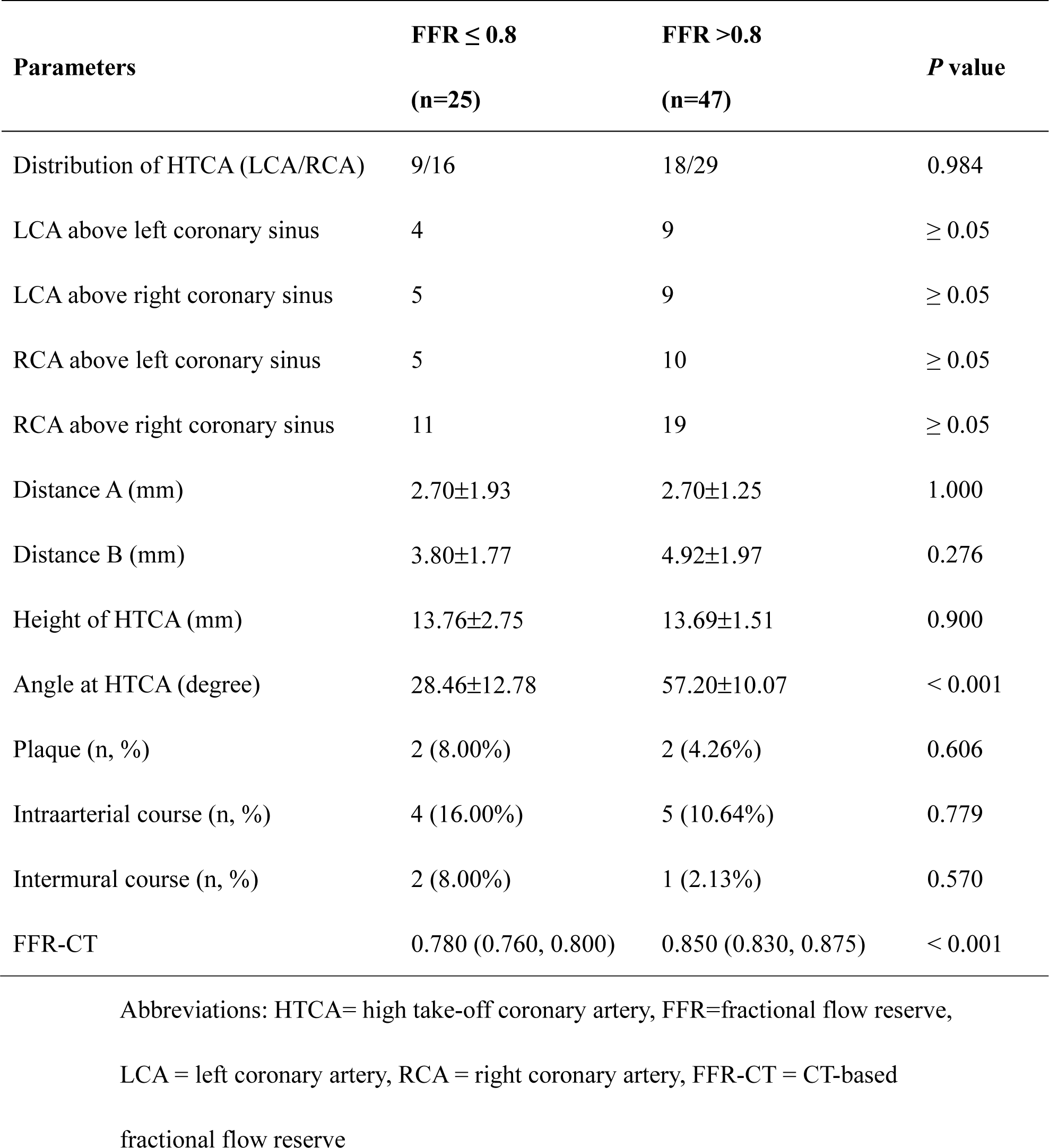
Comparison of anatomical features of the HTCA between two groups.

**Figure 2.**
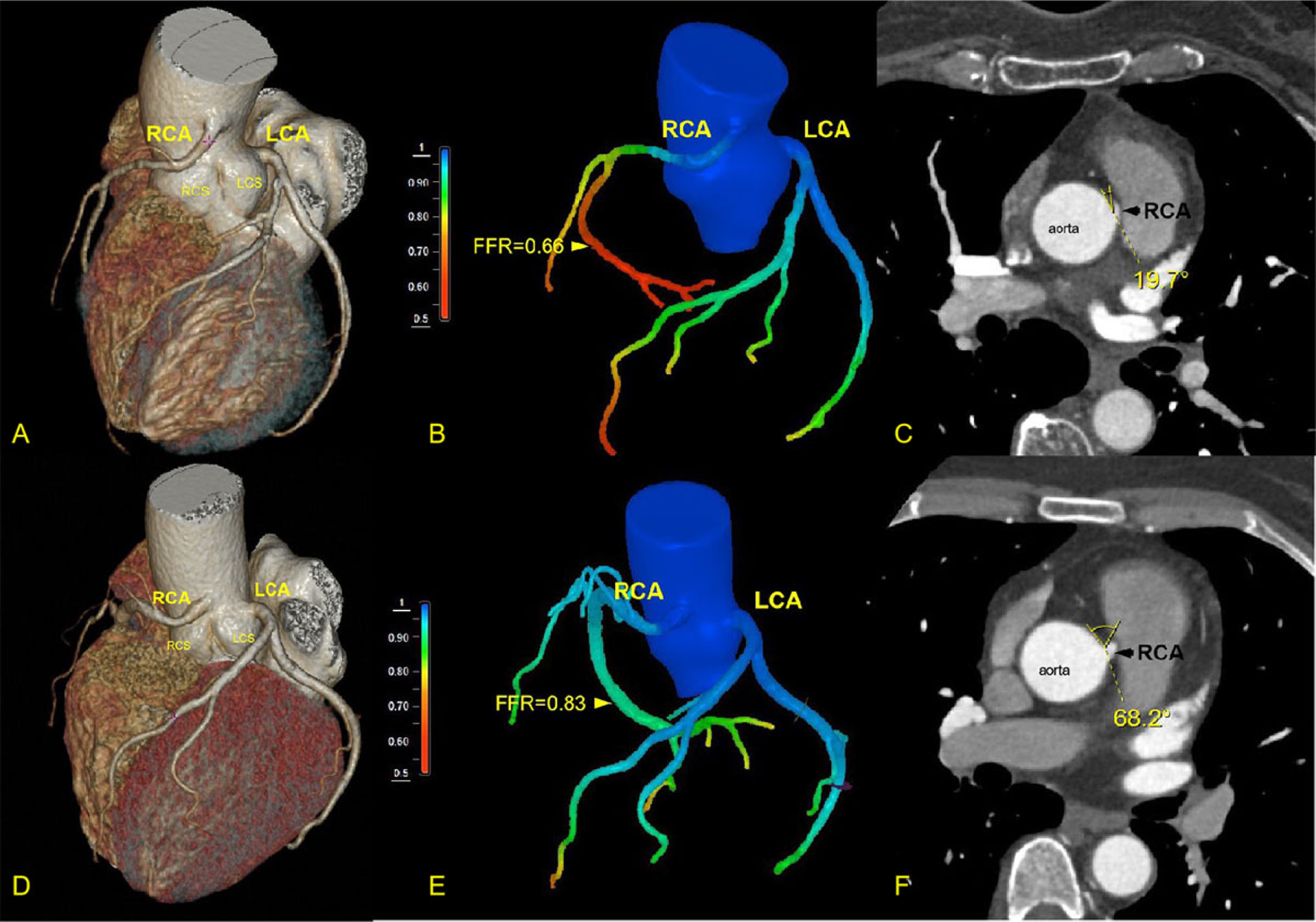
CCTA evaluation of the HTCA. Images A-C show a patient in the FFR ≤ 0.8 group: A) The VR image of CCTA showed high take-off RCA arising from the ascending aorta and above the left coronary sinus. B) The FFR-CT image showed hemodynamical change. The FFR-CT value of the distal of RCA was 0.66. C) The angle at the HTCA was measured at axial imaging and was 19.7 degrees. Images D-F show a patient in the FFR > 0.8 group: D) The VR image of CCTA showed high take-off RCA arising from the ascending aorta and above the right coronary sinus. B) The FFR-CT image showed hemodynamical change. The FFR-CT value of the distal of RCA was 0.83. C) The angle at the HTCA was measured at axial imaging and was 68.2 degrees. Abbreviations: CCTA = coronary computed tomography angiography, HTCA = high take-off coronary artery, FFR= fractional flow reserve, FFR-CT = CT-based fractional flow reserve, LCA = left coronary artery, RCA = right coronary artery

### Determination of independent predictive factors for abnormal hemodynamic changes in patients with HTCA

The above findings suggest that age, chest pain, palpitation, angle at the HTCA, and FFR-CT were significantly changed in both groups. These five factors were used to identify factors that can be used to predict significant hemodynamic changes in patients with HTCA. The factor of FFR-CT was first excluded for multicollinearity with FFR(VIF=73.811). Univariate analysis revealed that age, chest pain, palpitation, and angle at the HTCA were significant predictors of FFR ≤ 0.8 (Table 3). Multivariate logistic regression analysis revealed that the chest pain and the angle at the HTCA remained significant predictors (Table 3).

**Table 3.** Univariable and multivariable analysis: independent predictive factors for abnormal hemodynamic changes in patients with HTCA.

| Parameters | Univariable analysis |  |  | Multivariable analysis |  |  |
| --- | --- | --- | --- | --- | --- | --- |
|  | Odds ratio | 95% CI | P value | Odds ratio | 95% CI | P value |
| Age | 0.913 | 0.859-0.970 | 0.003 | 0.969 | 0.892-1.052 | 0.448 |
| chest pain | 6.385 | 2.685-15.187 | 0.000 | 3.201 | 1.113-9.203 | 0.031 |
| palpitation | 0.271 | 0.088-0.839 | 0.023 | 1.152 | 0.236-5.636 | 0.861 |
| Angel at HTCA | 0.867 | 0.816-0.923 | 0.000 | 0.891 | 0.832-0.955 | 0.001 |

Abbreviations: CI = confidence index, HTCA = high take-off coronary artery

Receiver operating characters curves analysis was then performed to identify the specificity and sensitivity of chest pain, angel at the HTCA, FFR-CT and combined indexes in evaluating the hemodynamic changes of patients with the HTCA. AUC, the best cutoff, sensitivity and specificity are listed in Table 4. According to ROC curve analysis (Fig. 3), the index of FFR-CT had significantly larger AUC than the index of chest pain (p=0.003). However, the AUC of the index of FFR-CT and angel at the HTCA was similar (p=0.059). Compared with combined index of angle at HTCA plus chest pain, FFR-CT had a larger AUC(p=0.000). The similar AUC was found between FFR-CT and the combined index of chest pain, angle at the HTCA plus FFR-CT (p=0.2359).

**Table 4.** Determination of predictive factors and FFR-CT for abnormal hemodynamic changes in patients with HTCA.

|  | **AUC**<br>**(95% CI)** | **Optimal cutoff**<br>**values** | **Sensitivity**<br>**(95% CI)** | **Specificity**<br>**(95% CI)** |
| --- | --- | --- | --- | --- |
| chest pain | 0.807<br>(0.697-0.801) | >1 | 87.10%<br>(70.2%-96.4%) | 58.54%<br>(42.1%-73.7%) |
| Angel at HTCA | 0.896<br>(0.802-0.956) | $\leq 37.3$ | 77.42%<br>(58.9%-90.4%) | 90.24%<br>(76.9%-97.3%) |
| FFR-CT | 0.958<br>(0.883-0.991) | $\leq 0.8$ | 87.10%<br>(70.2%-96.4%) | 95.12%<br>(83.5%-99.4%) |
| Chest pain+ Angle at HTCA | 0.700<br>(0.580-0.802) | --- | 74.19%<br>(55.4%-88.1%) | 63.41%<br>(46.9%-77.9%) |
| chest pain + angel at HTCA +<br>FFR-CT | 0.969<br>(0.899-0.996) | --- | 80.65%<br>(62.5%-92.5%) | 100%<br>(91.4%-100%) |

Abbreviations: HTCA = high take-off coronary artery, FFR-CT = CT-based fractional flow reserve, AUC = area under curve, CI = confidence interval

**Figure 3.**
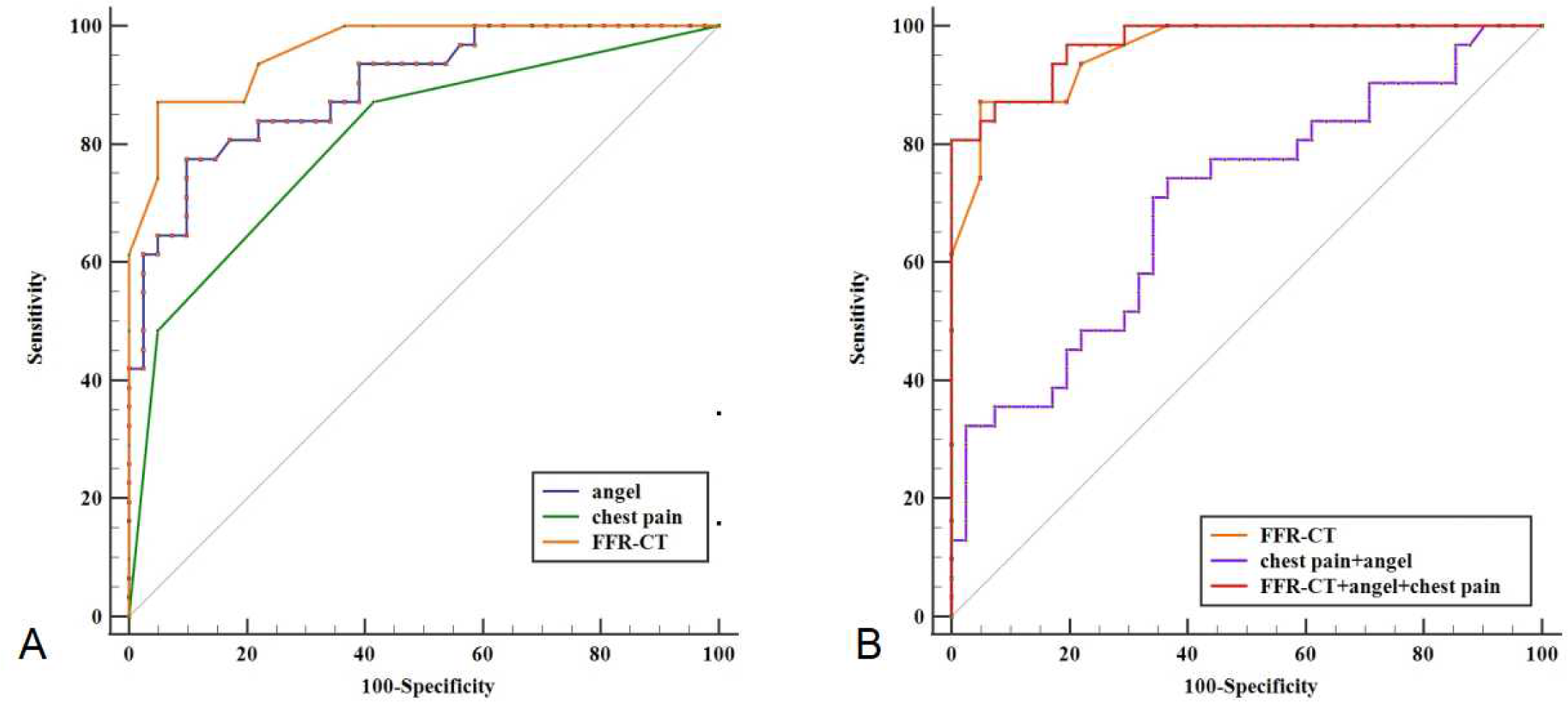
Comparison of ROC curves for significant hemodynamic change in HTCA. Image A comparison of AUC among the indexes of chest pain, angle at HTCA and FFR-CT. Image B comparison of AUC among combined indexes (FFR-CT plus angle at HTCA and chest pain, chest pain plus angle at the HTCA) with FFR-CT. Abbreviations: ROC = receiver operating characters, HTCA = high take-off coronary artery, AUC = area under curve

## Discussion

The major finding of present study was that FFR-CT, chest pain and the angle at the HTCA could be used to predict abnormal hemodynamic changes in patients with HTCA. FFR-CT had a superior sensitivity and specificity to predict abnormal hemodynamic changes of HTCA.

In this study, chest pain and the angle at the HTCA were independent factors in predicting significant hemodynamic changes of HTCA. These two independent factors contained clinical complications and anatomical factors. In clinical complications, chest pain is an independent risk factor. Corresponding with previous studies [14,17], chest pain is most common symptoms in HTCA. However, besides chest pain, palpitation, dyspnea and syncope were also reported in patients with adverse cardiac events in the previous study [17-20]. The possible reason for this result might be the difference of patient cohort. Our study focused on HTCA patients, not including other anomalies with HTCA. In addition, our study also found that preoperative ischemia and atypical symptoms should be paid more attention. In anatomical factors, height and angle at the HTCA were main anatomical features mentioned in previous study [2]. A height of 10 mm above the sinotubular junction was considered clinically significant [2]. In the study by Tang et al, patients with a height above the pulmonary artery were more likely to show abnormal FFR-CT values [21]. In our study, we found that the height was not an independent risk to predict significant hemodynamical change, which was consistent with many reported HTCA cases with adverse cardiac events. Compared with the height of the HTCA, the angle at the HTCA has been shown to have a definite association with adverse cardiac events. Virmani et al. autopsied patients of sudden death and found that an acute angle at the HTCA was more frequently observed in sudden death cases, suggesting that a dilated aortic root compressed the HTCA with an acute angle, causing acute obstruction [22]. Although some authors thought that the HTCA with an acute angle may be a harmless anomaly, angle at the HTCA was considered a worrisome feature in coronary anomaly [23-24]. In our study, the angle at the HTCA was an independent risk factor to predict significant hemodynamical changes. Considering different results of anatomical features in different study, anatomical features might play a complex role in HTCA.

However, in predicting significant hemodynamic change in patient with HTCA, FFR-CT played a more important role. First, the result of FFR-CT was consistent with FFR. The factor of FFR-CT was multicollinearity with FFR (VIF=73.811). FFR is a gold standard to assess functional status in coronary artery lesions currently [10]. However, in coronary anomaly, it was limited to be used for many factors, such as safety consideration [7]. In our study, patients were examined with FFR after CCTA for abnormal EKG. Compared to FFR, FFR-CT was calculated from standard CCTA and widely used in different hemodynamical status including patients with anomalous aortic origin of coronary artery. [25]. Second, FFR-CT had a good sensitivity and specificity to predict abnormal hemodynamic changes in patients with HTCA. In this study, the AUC of FFR-CT had significantly larger than the index of chest pain and combined index of angle at HTCA plus chest pain (p=0.003, p=0.000) and the similar AUC with the combined index of chest pain, angle at the HTCA plus FFR-CT (p=0.2359). It inferred that FFR-CT will be appreciated to evaluate the coronary anomaly, especial in potential adverse hemodynamical status. Although, FFR-CT was unusual used in coronary anomaly, our study would be a good try.

This study had some limitations. First, this was a retrospective study with a relatively small cohort size. Therefore, the bias may exist in our study. A large number of samples will be investigated to further confirm the accuracy of combined indexes. Second, we did not perform a follow-up study in this investigation, which limited the application of our findings in risk stratification of patients with HTCA. Thus, it gives us a room for improvement to revise our model to make a more accurate prediction.

## Conclusion

In conclusion, FFR-CT, chest pain and angle at HTCA based on CCTA played a great role to predict significant hemodynamic changes in patients with HTCA. FFR-CT had a superior diagnostic performance to predict abnormal hemodynamic changes in patients with HTCA. It may be helpful to choose an appropriate management of HTCA and to reduce adverse cardiac events of HTCA patients.

## Data Availability

All data generated or analysed during this study are included in this published article

## Non-standard Abbreviations and Acronyms

HTCA: High take-off coronary artery
RCA: the right coronary artery
LCA: the left coronary artery
ATCM: the automatic tube current modulation
FFR-CT: CT-based fractional flow reserve

## Acknowledgments

None.

## Author contributions

J.N., S.C. and C.G.perform material preparation, data collection and analysis. J.N., S.C. and C.G. wrote the main manuscript text. J.N., C.G. and P.W. contributed to the study conception and design. C.G.and P.W. commented on previous versions of the manuscript. All authors read and approved the final manuscript.

## References

1. Kim SY, Seo JB, Do KH, Heo JN, Lee JS, Song JW, Choe YH, Kim TH, Yong HS, Choi SI, et al. Coronary Artery Anomalies: Classification and ECG-gated Multi–Detector Row CT Findings with Angiographic Correlation. RadioGraphics. 2006; 26: 317–333.

2. Loukas M, Andall RG, Khan AZ, Patel K, Muresian H, Spicer DE, Tubbs RS. The clinical anatomy of high take-off coronary arteries. Clin Anat. 2016; 29:408–419.

3. Motamedi MH, Hemmat A, Kalani P, Rezaee MR, Safarnezhad S. High take-off of right coronary artery: an extremely rare case of RCA anomaly. J Card Surg. 2009; 24:343–345.

4. Nishida N, Hata Y, Kinoshita K. High take-off of the left main coronary artery at autopsy after sudden unexpected death in a male. Pathology. 2014; 46:361–364.

5. Kadakia J, Gupta M, Budoff MJ. Anomalous “High Take-Off” of the right coronary artery evaluated by coronary CT angiography. Cathet Cardiovasc Interv. 2013; 82: e765–768.

6. Wang SP, Jao YTFN, Han S-C. Acute coronary syndrome due to high aortocoronary junction of the right coronary artery: The value of multislice CT. Int J Card. 2008; 123: e59–61.

7. Driesen BW, Warmerdam EG, Sieswerda GT, Schoof PH, Meijboom FJ, Haas F, Stella PR, Kraaijeveld AO, Evens FCM, Doevendans PAFM, et al. Anomalous coronary artery originating from the opposite sinus of Valsalva (ACAOS), fractional flow reserve- and intravascular ultrasound-guided management in adult patients. Catheter Cardiovasc Interv. 2018;92:68–75.

8. Lee HJ, Hong YJ, Kim HY, Lee J, Hur J, Choi BW, Chang HJ, Nam JE, Choe KO, Kim YJ. Anomalous origin of the right coronary artery from the left coronary sinus with an interarterial course: subtypes and clinical importance. Radiology. 2012; 262:101–108.

9. Ko BS, Linde JJ, Ihdayhid AR, Norgaard BL, Kofoed KF, Sørgaard M, Adams D, Crossett M, Cameron JD, Seneviratne SK. Non-invasive CT-derived fractional flow reserve and static rest and stress CT myocardial perfusion imaging for detection of haemodynamically significant coronary stenosis. Int J Cardiovas Imaging. 2019; 35:2103–2112.

10. Min JK, Leipsic J, Pencina MJ, Berman DS, Koo BK, van Mieghem C, Erglis A, Lin FY, Dunning AM, Apruzzese P, et al. Diagnostic accuracy of fractional flow reserve from anatomic CT angiography. JAMA. 2012; 308: 1237–1245.

11. Ni J, Ge C, Li C, Ma J, Ren Y, Wang P. Computed tomographic coronary angiography combined computational fluid dynamics demonstrates the hemodynamic changes of a pathologic condition Vieussens’ arterial ring. Int J Cardiol. 2016; 223:398–400.

12. Yu M, Dai X, Deng J, Lu Z, Shen C, Zhang J. Diagnostic performance of perivascular fat attenuation index to predict hemodynamic significance of coronary stenosis: a preliminary coronary computed tomography angiography study. Eur Radiol. 2020; 30:673–681.

13. Ihdayhid AR, Norgaard BL, Gaur S, Leipsic J, Nerlekar N, Osawa K, Miyoshi T, Jensen JM, Kimura T, Shiomi H, et al. Prognostic Value and Risk Continuum of Noninvasive Fractional Flow Reserve Derived from Coronary CT Angiography. Radiology. 2019; 292:343–351.

14. Diao KY, Zhao Q, Gao Y, Shi K, Ma M, Xu HY, Guo YK, Yang ZG. Prognostic value of dual-source computed tomography (DSCT) angiography characteristics in anomalous coronary artery from the opposite sinus (ACAOS) patients: a large-scale retrospective study. BMC Cardiovasc Disord. 2020; 20: 25.

15. Tang CX, Liu CY, Lu MJ, Schoepf UJ, Tesche C, Bayer RR 2nd, Hudson HT Jr, Zhang XL, Li JH, Wang YN, et al. CT FFR for ischemia-specific CAD with a new computational fluid dynamics algorithm: a Chinese multicenter study. JACC Cardiovasc Imaging. 2020; 13:980–990.

16. Taylor CA, Fonte TA, Min JK. Computational fluid dynamics applied to cardiac computed tomography for noninvasive quantification of fractional flow reserve: scientific basis. J Am Coll Cardiol. 2013; 61: 2233–2241.

17. Padalino MA, Franchetti N, Hazekamp M. Surgery for anomalous aortic origin of coronary arteries: a multicentre study from the European Congenital Heart Surgeons Association. Eur J Cardiothorac Surg. 2019; 56:696–703.

18. Rahmouni K, Bernier PL. Current Management of Anomalous Aortic Origin of a Coronary Artery: A Pan-Canadian Survey. World J Pediatr Congenit Heart Surg. 2021; 12:387–393.

19. Brothers JA, Frommelt MA, Jaquiss RDB, Myerburg RJ, Fraser CD Jr, Tweddell JS. Expert consensus guidelines: Anomalous aortic origin of a coronary artery. J Thorac Cardiovasc Surg. 2017; 153:1440–1457.

20. Jegatheeswaran A, Alsoufi B. Anomalous aortic origin of a coronary artery: 2020 year in review. J Thorac Cardiovasc Surg. 2021; 162:353–359.

21. Tang CX, Lu MJ, Schoepf JU, Tesche C, Bauer M, Nance J, Griffith P, Lu GM, Zhang LJ. Coronary Computed Tomography Angiography-Derived Fractional Flow Reserve in Patients with Anomalous Origin of the Right Coronary Artery from the Left Coronary Sinus. Korean J Radiol. 2020; 21:192–202.

22. Virmani R, Chun PK, Goldstein RE, Robinowitz M, McAllister HA. Acute take-offs of the coronary arteries along the aortic wall and congenital coronary ostial valve-like ridges: association with sudden death. J Am Coll Cardiol. 1984; 3:766–771.

23. Nasis A, Machado C, Cameron JD, Troupis JM, Meredith IT, Seneviratne SK. Anatomic characteristics and outcome of adults with coronary arteries arising from an anomalous location detected with coronary computed tomography angiography. Int J Cardiovasc Imaging. 2015; 31:181–191.

24. Brothers JA, Gaynor JW, Jacobs JP, Caldarone C, Jegatheeswaran A, Jacobs ML; Anomalous Coronary Artery Working Group. The registry of anomalous aortic origin of the coronary artery of the congenital heart surgeons’ society. Cardiol Young. 2010; 20 suppl 3:50–58.

25. Ferrag W, Scalbert F, Adjedj J, Dupouy P, Ou P, Juliard JM, Farnoud R, Benadji AA, Du Fretay XH, Wijns W, et al. Role of FFR-CT for the evaluation of patients with anomalous aortic origin of coronary artery. JACC Cardiovasc Imaging. 2021;14:1074–1076.

